# Trends in long-term vaping among adults in England, 2013-2023

**DOI:** 10.1101/2023.12.21.23300376

**Authors:** Sarah E. Jackson, Harry Tattan-Birch, Lion Shahab, Jamie Brown

## Abstract

**Objectives:** To examine trends in long-term (>6 months) vaping among adults in England.

**Design:** Nationally-representative monthly cross-sectional survey.

**Setting:** England.

**Participants:** 179,725 adults (≥18y) surveyed between October 2013 and October 2023.

**Main outcome measures:** We used logistic regression to estimate time trends in the prevalence of long-term vaping, overall and by vaping frequency (daily/non-daily) and the main device type used (disposable/refillable/pod).

**Results:** The proportion of adults reporting long-term vaping increased non-linearly from 1.3% [95%CI 1.1-1.5%] in October 2013 to 10.0% [9.2-10.9%] in October 2023, with a particularly pronounced rise since 2021. This included an increase in long-term daily vaping, which rose from 0.6% [0.5-0.8%] to 6.7% [6.0-7.4%], respectively. The absolute increases in long-term vaping were most pronounced among ever smokers (current smokers: 4.8% [4.0-5.8%] to 23.1% [20.4-25.9%]; recent ex-smokers: 5.7% [3.4-9.2%] to 36.1% [27.6-45.4%]; long-term ex-smokers: 1.4% [1.0-1.9%] to 16.2% [14.2-18.4%]), but there was also an increase among never smokers (0.1% [0.0-0.2%] to 3.0% [2.3-3.8%]). Growth was also most pronounced in younger adults (e.g., reaching 22.7% [19.2-26.5%] of 18-year-olds vs. 4.3% [3.6-5.2%] of 65-year-olds), including among never smokers (reaching 16.1% [11.1-22.7%] among 18-year-olds vs. 0.3% [0.1-0.6%] of 65-year-olds). Up to March 2021, most long-term vapers mainly/exclusively used refillable e-cigarettes (2.5-3.3% of adults) and very few (0.1% of adults) used disposables. However, prevalence of long-term disposable vaping subsequently rose rapidly and by October 2023, similar proportions mainly/exclusively used refillable and disposable devices (4.6% [4.0-5.3%] and 4.9% [4.2-5.7%] of adults, respectively).

**Conclusions:** The prevalence of long-term (>6 months) vaping has increased substantially among adults in England over the past decade. Much of this increase in prevalence has occurred since 2021, coinciding with the rapid rise in popularity of disposable e-cigarettes. Half of long-term vapers now mainly or exclusively use disposable devices. The growth has been concentrated among ever smokers but there has also been an increase among never smokers, especially younger adults.

**What is already known on this topic:**

- Vaping prevalence has increased substantially in England since new disposable e-cigarettes became popular in mid-2021, particularly among young people.
- It is not clear how far this reflects an increase in experimental use versus long-term, regular use.
- In addition, little is known about how the types of products used by long-term vapers is changing over time.

**What this study adds:**

- There has been an exceptionally steep rise in long-term vaping among young adults since 2021, including among never smoking youth, and it does not yet show signs of stopping.
- Half of long-term vapers now mainly or exclusively use disposables, and most are using them every day.
- Therefore, urgent action is needed to curb the rise in vaping among young people and encourage long-term vapers to transition to less environmentally damaging products.

## Introduction

In England, the prevalence of vaping has risen in recent years, particularly among young adults.^1–3^ This has largely been attributed to the introduction of new disposable devices to the market.^4,5^ However, it is not clear how far this rise in prevalence reflects an increase in short-term, experimental use versus sustained, regular use. In addition, little is known about how the types of products used by long-term vapers – and how often they use them – is changing over time. Understanding this is important for gauging the likely public health and environmental impacts of this rise in vaping uptake and for informing policy decisions.

In the US, there was a rapid increase in the use of e-cigarettes in adolescents and young adults between 2015 and 2019, which sparked widespread public health concern.^6,7^ Further analyses of the data showed that much of the use was experimental,^8^ and there was little evidence of an overall increase in the population burden of nicotine dependence.^9^ Consistent with this pattern of use and following new regulations restricting the availability of e-cigarettes, there were subsequent declines in the prevalence of use in the same population.^10^ In the UK, there has been a sharp recent increase in vaping among young people.^3,4^ It is important to understand the extent to which this represents experimental or dependent use, and how this differs across subgroups of the population (e.g., by smoking status, age, gender, and socioeconomic position). One indicator of more dependent use is the prevalence of long-term vaping.

Long-term vaping could pose both benefits and harms to public health, depending on who is using the products, and what they would otherwise be doing. Transitioning to long-term vaping from cigarette smoking — the most harmful form of nicotine use — would be beneficial for people who are unable or unwilling to stop all nicotine use.^11^ Conversely, long-term vaping could lead to harm among those who would have never started smoking cigarettes.^11^ Long-term vaping with disposable e-cigarettes specifically may also have a substantial environmental impact:^12^ these products are designed to be single-use, so they generate more waste than rechargeable vaping products (i.e., refillable/pod devices).^13,14^

The Smoking Toolkit Study (a nationally-representative, cross-sectional survey) collects detailed data on vaping among adults in England each month. This study aimed to use data collected over the past decade to address the following research questions:

1. Among adults in England, how has the prevalence of long-term (>6 months) vaping changed between 2013 and 2023 – overall and by vaping frequency (daily/non-daily) and the main device type used (disposable/refillable/pod)?
2. Have changes in (i) any long-term vaping and (ii) long-term disposable vaping differed by smoking status, age, gender, or occupational social grade?

## Method

### Pre-registration

The study protocol and analysis plan were pre-registered on Open Science Framework (https://osf.io/n2785/). In addition to our pre-registered analyses, we added an unplanned analysis in which we modelled age-specific trends in long-term vaping among people who had never regularly smoked, to explore the extent to which the increase we observed among never smokers differed by age.

### Design

Data were drawn from the ongoing Smoking Toolkit Study, a monthly cross-sectional survey of a representative sample of adults in England.^15^ The study uses a hybrid of random probability and simple quota sampling to select a new sample of approximately 1,700 adults each month. Comparisons with sales data and other national surveys indicate that key variables including sociodemographic characteristics, smoking prevalence, and cigarette consumption are nationally representative.^15,16^

Data were initially collected through face-to-face computer-assisted interviews. However, social distancing restrictions under the Covid-19 pandemic meant no data were collected in March 2020 and data from April 2020 onwards were collected via telephone. The telephone-based data collection used broadly the same combination of random location and quota sampling, and weighting approach as the face-to-face interviews and comparisons of the two data collection modalities indicate good comparability.^17–19^

For this study, we used data collected from participants surveyed between October 2013 (the first wave to assess vaping among all adults) and October 2023 (the most recent data at the time of analysis). Since April 2022, vaping duration and device characteristics have only been assessed quarterly due to availability of competitive research funding. Waves in which vaping duration was not assessed (May/June/August/September/November/December 2022, February/March/May/August/September 2023) were excluded. Data on main device type used have only been collected since July 2016, so trends in this outcome were limited to the period from July 2016 to October 2023. Because data were not collected from 16- and 17-year-olds between April 2020 and December 2021, we restricted our sample to those aged ≥18 for consistency across the time series.

### Measures

***Long-term vaping*** was defined as current vaping for a period of more than 6 months. Current vaping as assessed within several questions asking about use of a range of nicotine products. Current smokers were asked ‘Do you regularly use any of the following in situations when you are not allowed to smoke?’; current smokers and those who had quit in the past year were asked ‘Can I check, are you using any of the following either to help you stop smoking, to help you cut down or for any other reason at all?’; and non-smokers were asked ‘Can I check, are you using any of the following?’. Those who reported using an e-cigarette in response to any of these questions were considered current vapers. Current vapers were then asked: ‘How long have you been using this nicotine replacement product or these products for?’ Response options were:

a. Less than one week
b. One to six weeks
c. More than six weeks up to twelve weeks
d. More than twelve weeks and up to 26 weeks
e. More than 26 weeks and up to 52 weeks
f. More than 52 weeks

Participants who reported having been vaping for more than 6 months (responses *e* and *f*) were considered long-term vapers.^20^ This definition was used to indicate possible dependent use.

***Vaping frequency*** was assessed by asking vapers: ‘How many times per day on average do you use your nicotine replacement product or products?’ Those who responded that they do not use it every day were asked whether they use it at least weekly. Those who reported use at least once a day were considered to be vaping daily and those who reported use less than once a day were considered to be vaping non-daily.

***Main device type used*** was assessed from July 2016 onwards by asking vapers: ‘Which of the following do you mainly use…?’ Response options were:

- *Disposable* – ‘A disposable e-cigarette or vaping device (non-rechargeable)’
- *Refillable* – ‘An e-cigarette or vaping device with a tank that you refill with liquids (rechargeable)’ or ‘A modular system that you refill with liquids (you use your own combination of separate devices: batteries, atomizers, etc.)’
- *Pod* – ‘An e-cigarette or vaping device that uses replaceable pre-filled cartridges (rechargeable)’

***Smoking status*** was assessed by asking participants which of the following best applies to them:

a. I smoke cigarettes (including hand-rolled) every day
b. I smoke cigarettes (including hand-rolled), but not every day
c. I do not smoke cigarettes at all, but I do smoke tobacco of some kind (e.g., pipe, cigar, or shisha)
d. I have stopped smoking completely in the last year
e. I stopped smoking completely more than a year ago
f. I have never been a smoker (i.e., smoked for a year or more)

Those who responded *a-c* were considered current smokers. Those who responded *d* were considered recent ex-smokers. Those who responded *e* were considered long-term ex-smokers. Those who responded *f* were considered never-smokers.

***Age*** was modelled as a continuous variable using restricted cubic splines (see *statistical analysis* section). We also provided descriptive data by age group (18-24/25-34/35-44/45-54/55-64/≥65).

***Gender*** was self-reported as man or woman. In more recent waves, participants have also had the option to describe their gender in another way; those who identified in another way were excluded from analyses by gender due to low numbers.

***Occupational social grade*** was categorised as ABC1 (includes managerial, professional, and upper supervisory occupations) and C2DE (includes manual routine, semi-routine, lower supervisory, and long-term unemployed).

### Statistical analysis

Data were analysed in R v.4.2.1. The Smoking Toolkit Study uses raking to weight the sample to match the population in England. This profile is determined each month by combining data from the UK Census, the Office for National Statistics mid-year estimates, and the annual National Readership Survey.^15^ The following analyses used weighted data. Missing cases were excluded on a per-analysis basis.

### RQ1. Trends in long-term vaping among adults

Among all adults, we reported the prevalence (with 95% confidence interval [CI]) of the following by survey year:

i. Long-term vaping (i.e., vaping for >6 months)
ii. Long-term daily vaping (i.e., vaping for >6 months and currently vaping daily)
iii. Long-term non-daily vaping (i.e., vaping for >6 months and currently vaping non-daily)
iv. Long-term disposable vaping (i.e., vaping for >6 months and currently mainly/exclusively using disposable e-cigarettes)
v. Long-term refillable vaping (i.e., vaping for >6 months and currently mainly/exclusively using refillable e-cigarettes)
vi. Long-term pod vaping (i.e., vaping for >6 months and currently mainly/exclusively using pod e-cigarettes)

Trends in these long-term vaping outcomes over the study period were analysed using logistic regression with time (survey month) modelled using restricted cubic splines with five knots (sufficient to accurately model trends across years without overfitting). This allowed for flexible and non-linear changes over time, while avoiding categorisation.

### RQ2. Differences by smoking status, age, gender, and occupational social grade

To explore moderation of trends in (i) any long-term vaping and (ii) long-term disposable vaping, by smoking status, age, gender, and occupational social grade, we repeated these models including the interaction between the moderator of interest and time – thus allowing for time trends to differ across sub-groups. Each of the interactions was tested in a separate model. Age was modelled using restricted cubic splines with three knots (placed at the 5, 50, and 95% quantiles), to allow for a non-linear relationship between age and long-term vaping.

In an unplanned analysis, we then repeated the model testing the interaction between age and time among participants who had never regularly smoked.

We used predicted estimates from these models to plot the prevalence of long-term vaping over the study period, among all adults and within each subgroup of interest. As age was modelled continuously, we displayed estimates for six specific ages (18-, 25-, 35-, 45-, 55-, and 65-year-olds) to illustrate how trends differ across ages. Note that the model used to derive these estimates included data from participants of all ages, not only those aged exactly 18, 25, 35, 45, 55, or 65 years.

### Patient and public involvement

The wider Smoking Toolkit Study is discussed several times a year with a diverse PPI group and the authors regularly attend and present at meetings at which patients and the public are included. Interaction and discussion at these events help shape the broad research priorities and questions. There is also a mechanism for generalised input from the wider public: each month interviewers seek feedback on the questions from all respondents, who are representative of the English population. This feedback is limited and usually relates to understanding of questions and item options. No patients or members of the public were involved in setting the research questions or the outcome measures, nor were they involved in the design and implementation of this specific study. Results will be disseminated to stakeholders in relevant public health bodies and charities.

## Results

A total of 197,266 (unweighted) adults aged ≥18 years in England were surveyed between October 2013 and October 2023. We excluded 17,541 surveyed in months in which vaping duration was not assessed, resulting in a final analytic sample of 179,725 participants. Of these, 125,751 were surveyed between July 2016 and October 2023 and provided data for analyses by main device type used. Characteristics of the whole analysed sample and the device type subsample are shown in **Table S1**.

### Trends in long-term vaping among adults

Across the study period, the proportion of adults reporting long-term vaping increased from 1.3% to 10.0% (**Table 1**). The increase over time was non-linear: prevalence increased from 1.3% to 3.3% between October 2013 and July 2017, was stable at 3.3% between July 2017 and August 2019, then increased again – with a particularly sharp rise since late 2021 – reaching 10.0% by October 2023 (**Figure 1**).

**Table 1.**
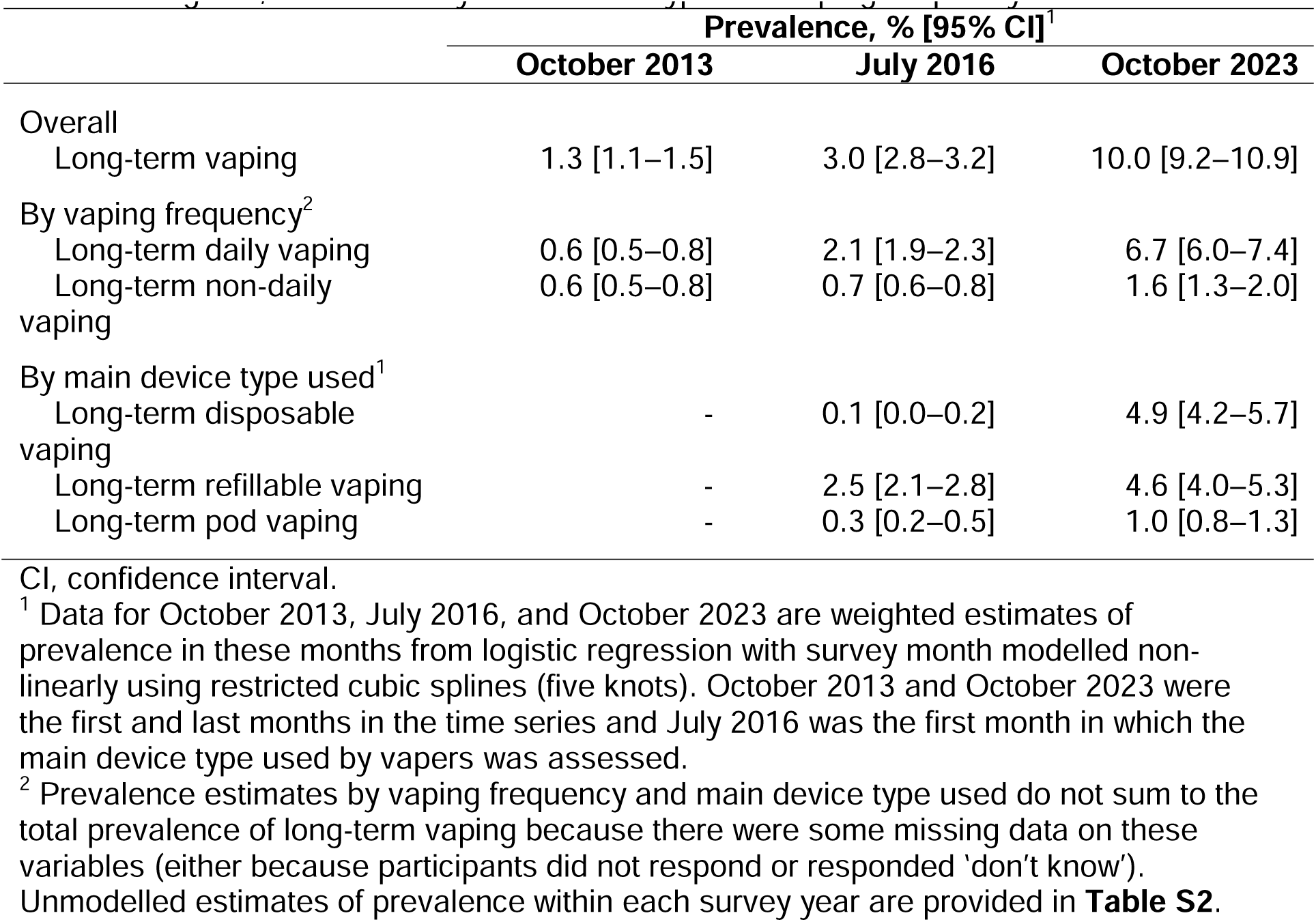
Modelled estimates of changes in the prevalence of long-term vaping among adults in England, overall and by main device type and vaping frequency

**Figure 1.**
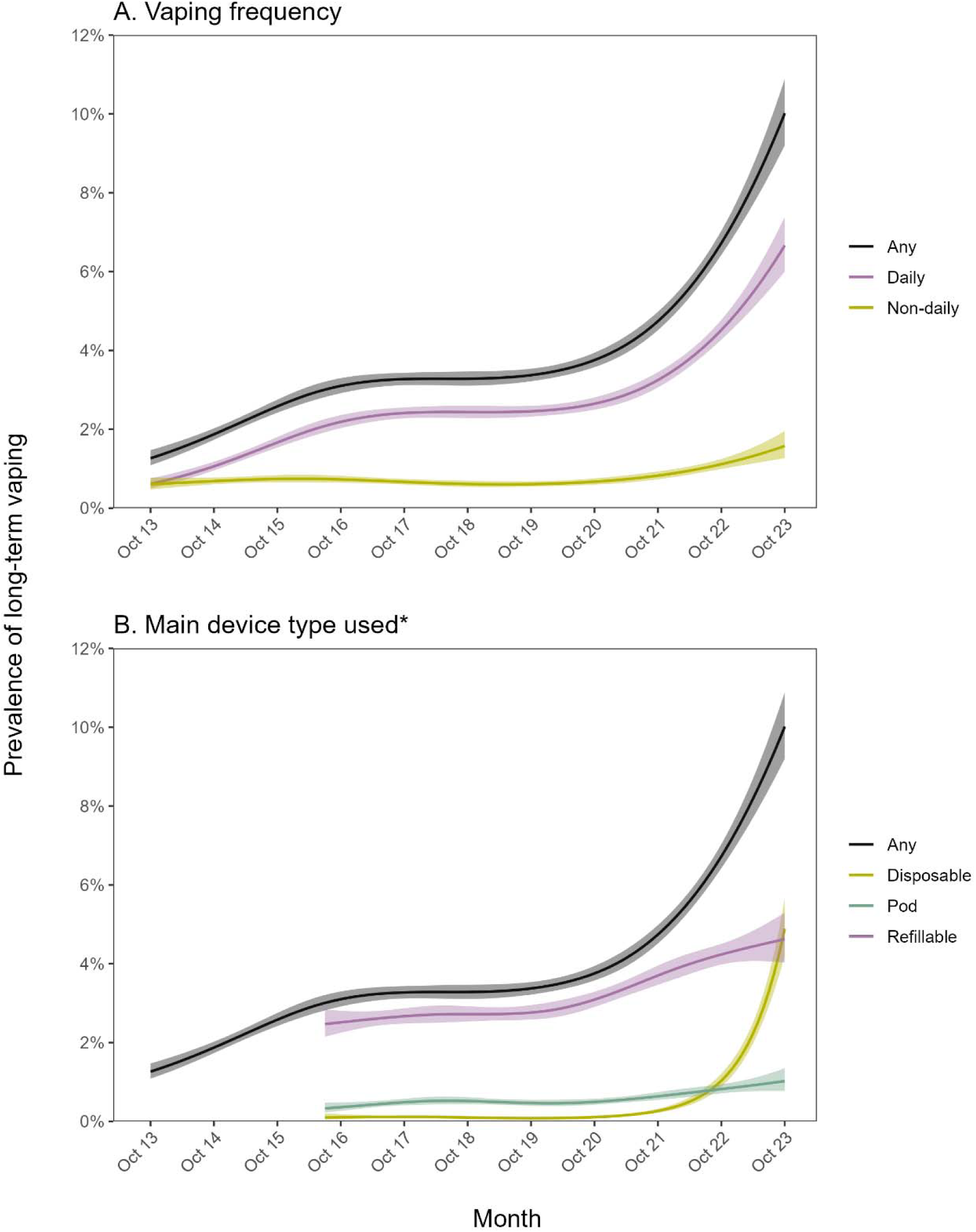
Trends in long-term vaping among adults in England, October 2013 to October 2023. Panels show trends in the prevalence of long-term (>6 months) vaping among adults in England, overall and by (A) vaping frequency (daily/non-daily) and (B) the main device type used (disposable/refillable/pod). Lines represent modelled weighted prevalence by monthly survey wave, modelled non-linearly using restricted cubic splines (five knots). Shaded bands represent 95% confidence intervals. *The main device type used was not assessed before July 2016. Note that prevalence estimates by vaping frequency and main device type used do not sum to the total prevalence of long-term vaping because there were some missing data on these variables (either because participants did not respond or responded ‘don’t know’).

There was a greater increase in long-term daily vaping than long-term non-daily vaping over time. In October 2013, equal proportions of long-term vapers reported vaping daily and non-daily (0.6% and 0.6% of adults, respectively; **Table 1**). The trend in long-term daily vaping mirrored the trend in any long-term daily vaping, increasing to 6.8% of adults by October 2023 (**Figure 1A**). Meanwhile, the prevalence of long-term non-daily vaping remained relatively stable (between 0.6% and 0.7%) up to May 2021, then increased to 1.6% by October 2023 (**Figure 1A**).

Trends also differed according to the main device type used. In July 2016, when device type was first assessed, most long-term vapers mainly/exclusively used refillable e-cigarettes (2.5% of adults) and very few mainly/exclusively used disposables (0.1% of adults; **Table 1**). By October 2023, similar proportions mainly/exclusively used refillable and disposable devices (4.6% and 4.9% of adults, respectively; **Table 1**). The prevalence of long-term disposable vaping was very low (0.1%) between October 2013 and March 2021, then increased rapidly to 4.9% by October 2023 (**Figure 1B**). The prevalence of long-term refillable vaping increased from 2.5% to 2.7% between October 2013 and September 2017, was stable at 2.7% up to September 2019, then increased to 4.6% by October 2023 (**Figure 1B**). The prevalence of long-term pod vaping was roughly stable (between 0.3% and 0.5%) from October 2013 to March 2021, then increased steadily to 1.0% by October 2023 (**Figure 1B**).

### Differences by smoking status, age, gender, and occupational social grade

The increase in long-term vaping occurred predominantly among current and ex-smokers, but there was also a rise among never smokers in more recent years (from <0.5% up to March 2021 to 3.0% by October 2023; **Table 2**, Figure 2A).

**Table 2.** Modelled estimates of changes in the prevalence of long-term vaping and long-term disposable vaping within subgroups of adults in England

|  | Any long-term vaping,<br>% [95% CI]<sup>1</sup> |  | Long-term disposable<br>vaping,<br>% [95% CI]<sup>1</sup> |  |
| --- | --- | --- | --- | --- |
|  | October<br>2013 | October 2023 | July 2016 | October 2023 |
| Smoking status |  |  |  |  |
| Never smoker | 0.1 [0.0–0.2] | 3.0 [2.3–3.8] | 0.0 [0.0–0.3] | 1.8 [1.2–2.5] |
| Long-term (≥1y) ex-smoker | 1.4 [1.0–1.9] | 16.2 [14.2–18.4] | 0.1 [0.0–0.5] | 4.4 [3.1–6.0] |
| Recent (<1y) ex-smoker | 5.7 [3.4–9.2] | 36.1 [27.6–45.4] | 0.3 [0.0–3.9] | 24.9 [15.9–36.8] |
| Current smoker | 4.8 [4.0–5.8] | 23.1 [20.4–25.9] | 0.4 [0.2–0.9] | 13.6 [11.1–16.6] |
| Age (years)<sup>2</sup> |  |  |  |  |
| 18 | 0.7 [0.4–1.0] | 22.7 [19.2–26.5] | 0.0 [0.0–0.2] | 19.1 [14.8–24.4] |
| 25 | 1.0 [0.7–1.3] | 18.6 [16.7–20.8] | 0.0 [0.0–0.2] | 12.2 [10.2–14.6] |
| 35 | 1.5 [1.2–1.8] | 13.8 [12.5–15.2] | 0.1 [0.0–0.2] | 6.3 [5.1–7.7] |
| 45 | 1.9 [1.5–2.3] | 9.9 [8.6–11.2] | 0.2 [0.1–0.4] | 3.2 [2.4–4.3] |
| 55 | 1.7 [1.4–2.1] | 6.7 [5.8–7.7] | 0.2 [0.1–0.6] | 1.8 [1.3–2.4] |
| 65 | 1.1 [0.9–1.4] | 4.3 [3.6–5.2] | 0.2 [0.1–0.4] | 1.0 [0.7–1.6] |
| Gender |  |  |  |  |
| Men | 1.2 [1.0–1.5] | 10.1 [8.9–11.3] | 0.1 [0.0–0.2] | 5.1 [4.1–6.3] |
| Women | 1.3 [1.1–1.6] | 9.9 [8.8–11.2] | 0.2 [0.1–0.4] | 4.7 [3.8–5.8] |
| Occupational social grade |  |  |  |  |
| ABC1 (more advantaged) | 1.0 [0.8–1.2] | 8.4 [7.6–9.3] | 0.1 [0.0–0.3] | 4.1 [3.4–5.0] |
| C2DE (less advantaged) | 1.6 [1.3–2.0] | 12.1 [10.6–13.7] | 0.1 [0.0–0.3] | 5.8 [4.6–7.3] |

CI, confidence interval.
<sup>1</sup> Data for October 2013, July 2016, and October 2023 are weighted estimates of prevalence in these months from logistic regression with survey month modelled non-linearly using restricted cubic splines (five knots). October 2013 and October 2023 were the first and last months in the time series and July 2016 was the first month in which the main device type used by vapers was assessed.
<sup>2</sup> Note that the model used to derive these estimates included data from participants of all ages, not only those who were aged exactly 18, 25, 35, 45, 55, or 65 years.

**Figure 2.**
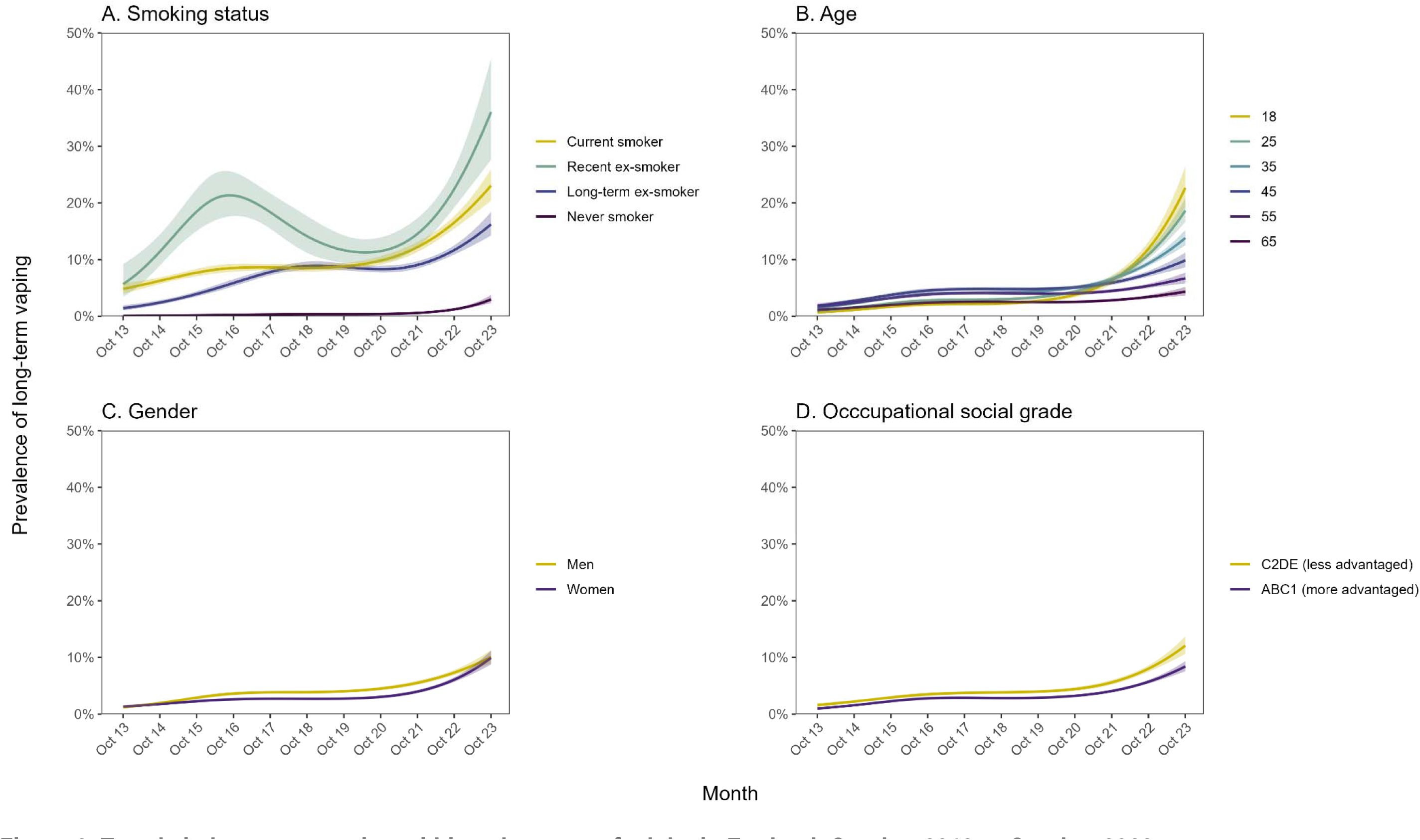
Trends in long-term vaping within subgroups of adults in England, October 2013 to October 2023. Panels show trends in the prevalence of long-term (>6 months) vaping among adults in England by (A) smoking status, (B) age, (C) gender, and (D) occupational social grade. Lines represent modelled weighted prevalence by monthly survey wave, modelled non-linearly using restricted cubic splines (five knots). Shaded bands represent 95% confidence intervals.

Changes in long-term vaping were similar across ages up to 2019, but prevalence then increased more rapidly among younger than older adults (Figure 2B), resulting in a strong age gradient in long-term vaping (e.g., reaching 22.7% among 18-year-olds vs. 4.3% among 65-year-olds by October 2023; **Table 2**). There was also a similar age gradient among never smokers (e.g., reaching 16.1% among 18-year-olds vs. 0.3% among 65-year-olds; **Table S3**, **Figure S1**).

The prevalence of long-term vaping initially rose slightly more quickly among men and, as a result, was significantly higher among men than women between June 2015 and December 2022 (Figure 2C). However, prevalence then increased more quickly among women than men since late 2021, closing this gap (Figure 2C). As of October 2023, there was no difference in the prevalence of long-term vaping between men (10.1%) and women (9.9%; **Table 2**).

The prevalence of long-term vaping was consistently higher among those from less vs. more advantaged social grades, but time trends were similar (Figure 2D).

Recent changes in long-term disposable vaping by smoking status, age, gender, and occupational social grade followed similar patterns to those observed for changes in any long-term vaping (**Table 2**, Figure 3).

**Figure 3.**
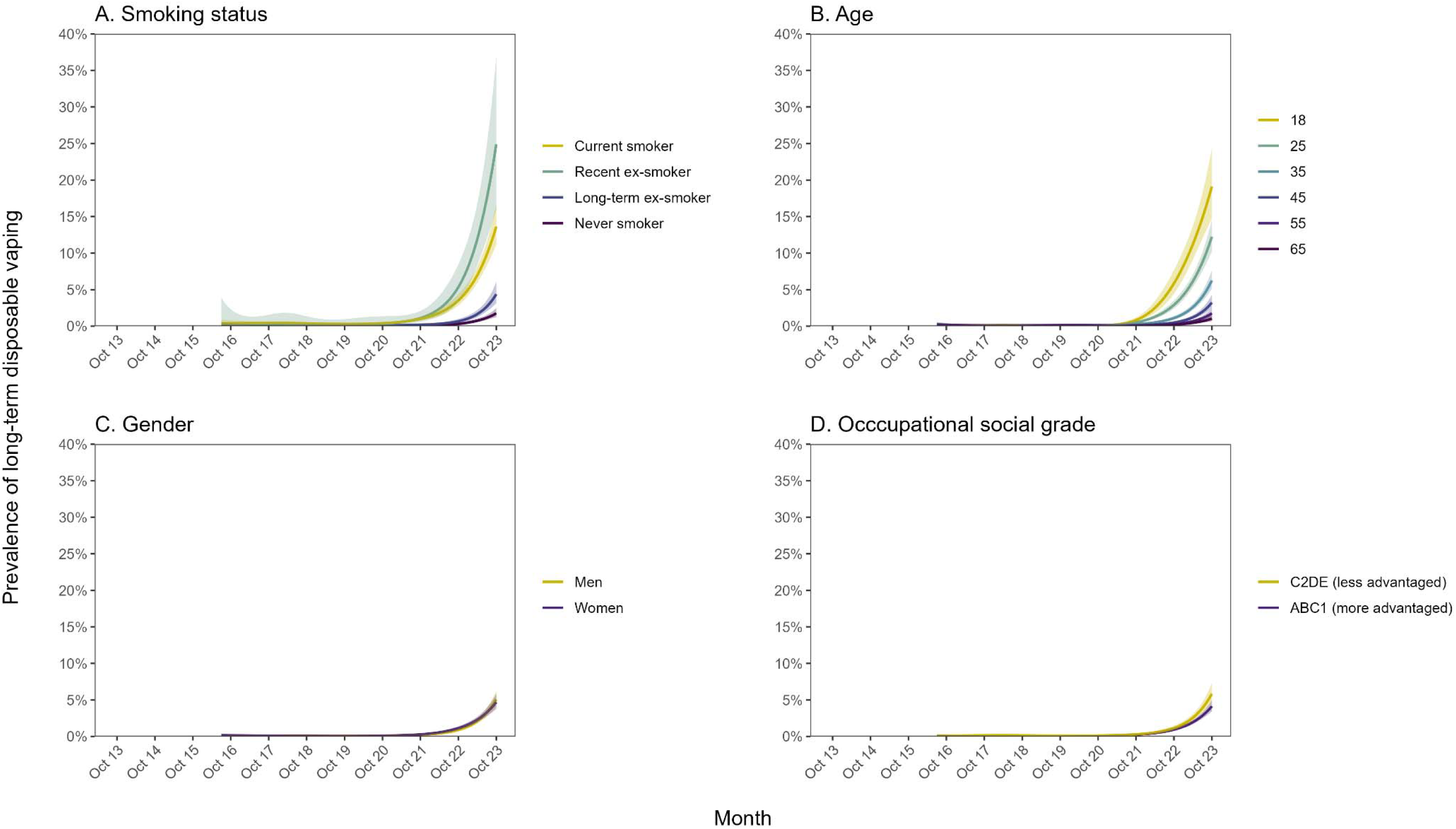
Trends in long-term disposable vaping within subgroups of adults in England, July 2016 to October 2023. Panels show trends in the proportion of adults in England reporting long-term (>6 months) vaping and currently mainly using disposable e-cigarettes, by (A) smoking status, (B) age, (C) gender, and (D) occupational social grade. Lines represent modelled weighted prevalence by monthly survey wave, modelled non-linearly using restricted cubic splines (five knots). Shaded bands represent 95% confidence intervals.

## Discussion

In England, the prevalence of long-term (>6 months) vaping has increased substantially over the past decade. In October 2013, around one in 80 adults were long-term vapers; by October 2023, this number had risen to one in 10. This increase was non-linear, with a rapid and unprecedented rise since 2021. The rise in long-term vaping was largely driven by an increase in long-term daily vaping. The absolute increases in long-term vaping were most pronounced among ever smokers, but there was also an increase among people who had never regularly smoked. Growth was also most pronounced in younger adults, including among never smokers. Up to March 2021, most long-term vapers mainly/exclusively used refillable e-cigarettes and very few used disposables. However, the prevalence of long-term disposable vaping subsequently rose rapidly and by October 2023, similar proportions mainly/exclusively used refillable and disposable devices. The prevalence of long-term pod vaping increased between March 2021 and October 2023, but remained relatively rare.

The timing of the sharp rise in long-term vaping coincides with the rising popularity of new disposable e-cigarettes since Spring 2021.^4,5^ This suggests that the recent increase in vaping among adults — particularly young adults — in England does not just reflect an increase in experimental use, but rather that a substantial number of those taking up vaping are going on to vape long-term. In addition, while there were small increases in long-term vaping with refillable and pod e-cigarettes in recent years, the increase in long-term vaping with disposable e-cigarettes was substantially larger. This indicates many vapers are now continuing to use disposables over the long-term, rather than transitioning to rechargeable devices.

We observed notable differences in trends in long-term vaping by age and smoking status. The recent and rapid rise in long-term vaping followed a clear age gradient that mirrored the pattern we have seen for the prevalence of current disposable vaping over this period.^4,5^ The rise occurred predominantly among current and ex-smokers, but there was also an increase in long-term vaping among those who had never regularly smoked. This is not necessarily a cause for concern if vaping is diverting people who would have otherwise smoked towards a less harmful nicotine product.

However, if these people would *not* have otherwise taken up smoking, then taking up vaping as a regular habit will expose them to greater harm than if they had neither vaped nor smoked.^11^ With an increasing proportion of people using e-cigarettes, it becomes increasingly unlikely that all would have used cigarettes instead given declining trends in the prevalence of cigarette smoking.^21^ Recent data indicate there has been a rise in the proportion of young adults using inhaled nicotine (i.e., vaping or smoking) since disposable e-cigarettes have become popular.^22^

We note that our definition of long-term vaping as use of e-cigarettes for a period of at least 6 months was an operational definition to indicate possible dependent use. It is not necessarily indicative of a length of time at which we would expect substantial harms from vaping to have already accumulated. In discussions about the risks of e-cigarettes, a key concern is that the harms of long-term use are not yet fully understood – in this context, long-term use refers to decades rather than the much shorter duration of use our analysis has focused on. Even the well-established harmful effects of tobacco smoking on health outcomes only become apparent after years and decades of use, rather than months. Existing evidence suggests the long-term health risks of vaping are likely to be substantially lower than the risks of smoking.^11^

Our findings raise two key issues with implications for policy. First, disposable e-cigarettes appear to be attracting young adults to establish long-term vaping use. There has been an exceptionally steep rise in long-term vaping among young adults since 2021 and it does not yet show signs of stopping. This adds weight to calls for tighter regulation of vaping products to reduce their appeal to young people^23^ and highlights the urgency for this action. Disposables may appeal to young people for a variety of reasons, including their low cost, sleek design and branding, attractive in-store displays, and ease of access.^23–25^ Policy options that may help to reduce young people’s interest in vaping, without detracting from the effectiveness of e-cigarettes as a smoking cessation aid,^26,27^ include applying an excise tax to disposables to increase the price and make them less affordable, restricting branding across all e-cigarette products and packaging, and restricting the visibility and promotion of e-cigarettes in shops.^23^

The second issue is that as of October 2023, half of long-term vapers are mainly or exclusively using disposables (and most are using them every day), which has a substantial environmental impact. A typical disposable e-cigarette is designed to be single-use and contains plastic, rubber, copper, and a lithium battery. While some parts (e.g., the battery) can be widely recycled, other parts (e.g., rubber) cannot, and devices are not generally designed to be taken apart easily. If they are not disposed of correctly, they can potentially release plastic, electronical, and hazardous chemical waste into the environment.^12^ There have been calls for an outright ban on disposable e-cigarettes.^28–30^ However, these products offer advantages for certain groups (e.g., those with severe mental illness, who may find them easier to use^31,32^) and in certain situations (e.g., for short-term use when a person has forgotten their rechargeable e-cigarette or it has run out of battery while out). Raising the upfront cost of disposable e-cigarettes to the same level as the cheapest reusable e-cigarettes (e.g., via an excise tax), thus making reusable e-cigarettes a more cost-effective alternative, may be effective in encouraging long-term vapers to transition to less environmentally damaging products over time,^23^ while allowing disposables to remain available to those who need them more.

Strengths of this study include the large, representative sample and repeated monthly assessment over a 10-year period. There were also limitations. The main device type used by vapers was not assessed before July 2016, so we were unable to model trends in long-term vaping by device type across the entire period. In addition, the items assessing vaping frequency and main device type only captured current behaviour, so our data do not offer insight into how these variables had changed within individuals with increasing duration of vaping. Finally, our definition of never smoking was not having regularly smoked for a year or more, so this group will include some people who have smoked regularly, but for less than a year. However, up to 2020, this measure indicated there was very low prevalence of long-term vaping among never smokers in the youngest age groups and much higher prevalence among ever smokers. Given the considerable change in trends we have observed, it seems unlikely that this limitation could explain the dramatic rise in long-term vaping. Moreover, even if everyone who reported long-term vaping had a history of smoking (whether or not they disclosed it in response to the item assessing smoking status), this would represent a serious departure from recent trends in smoking that warrants attention.

In conclusion, the prevalence of long-term (>6 months) vaping has increased substantially among adults in England over the past decade. Much of this increase in prevalence has occurred since 2021, coinciding with the rapid rise in popularity of disposable e-cigarettes. Half of long-term vapers now mainly or exclusively use disposable devices. The growth has been concentrated among ever smokers but there has also been an increase among never smokers, especially younger adults.

## Data Availability

The data used for these analyses are available on Open Science Framework (https://osf.io/n2785/), with age provided in bands to preserve participant anonymity.

## Declarations

### Acknowledgments

We thank Action on Smoking and Health for their comments on a late draft of the manuscript.

## Ethics approval

Ethical approval for the STS was granted originally by the UCL Ethics Committee (ID 0498/001). The data are not collected by UCL and are anonymized when received by UCL.

## Competing interests

JB has received unrestricted research funding from Pfizer and J&J, who manufacture smoking cessation medications. LS has received honoraria for talks, unrestricted research grants and travel expenses to attend meetings and workshops from manufactures of smoking cessation medications (Pfizer; J&J), and has acted as paid reviewer for grant awarding bodies and as a paid consultant for health care companies. All authors declare no financial links with tobacco companies, e-cigarette manufacturers, or their representatives.

## Funding

Cancer Research UK (PRCRPG-Nov21\100002) funded the Smoking Toolkit Study data collection and salary for SJ and HTB.

The funder played no role in the study design; in the collection, analysis, and interpretation of data; in the writing of the report; and in the decision to submit the article for publication. The authors are independent from the funders and that all authors had full access to all of the data (including statistical reports and tables) in the study and can take responsibility for the integrity of the data and the accuracy of the data analysis.

For the purpose of Open Access, the author has applied a CC BY public copyright licence to any Author Accepted Manuscript version arising from this submission.

## Notes

### Clinical Protocols

https://osf.io/n2785/

### Funding Statement

This study was funded by Cancer Research UK (PRCRPG-Nov21\100002).

