## Supplementary material for "Trends in long-term vaping among adults in England, 2013-2023": Table S1

##### Table S1. Weighted sample characteristics

|  | **Adults surveyed October 2013 – October 2023**  **(*n*^1^=179,725)** | **Adults surveyed October 2013 – October 2023**  **(*n*^1^=125,751)** |
| --- | --- | --- |
| Smoking status |  |  |
| Never smoker | 62.0% | 62.0% |
| Long-term (≥1y) ex-smoker | 18.8% | 19.3% |
| Recent (<1y) ex-smoker | 1.6% | 1.7% |
| Current smoker | 17.6% | 17.0% |
| Missing, *n***^1^** | 458 | 426 |
| Age (years) |  |  |
| Mean (SD) | 47.9 (18.6) | 48.1 (18.7) |
| 16-24 | 12.4% | 12.2% |
| 25-34 | 17.2% | 17.2% |
| 35-44 | 16.4% | 16.1% |
| 45-54 | 17.3% | 17.2% |
| 55-64 | 14.6% | 14.7% |
| ≥65 | 22.1% | 22.6% |
| Gender |  |  |
| Men | 48.9% | 48.9% |
| Women | 50.9% | 50.8% |
| Other | 0.2% | 0.3% |
| Missing, *n***^1^** | 66 | 66 |
| Occupational social grade |  |  |
| ABC1 (more advantaged) | 54.4% | 55.7% |
| C2DE (less advantaged) | 44.6% | 44.3% |

^1^ Unweighted sample size.

Note: Data are shown as weighted column percentages, unless otherwise specified. There were some missing data (unweighted *n*s indicated in the table); valid percentages are shown for ease of interpretation.

##### Table S2. Prevalence of long-term vaping by year among adults in England, 2013/14 to 2022/23

|  | **Prevalence, % [95% CI]^1^** | | | | | | | | | |
| --- | --- | --- | --- | --- | --- | --- | --- | --- | --- | --- |
|  | **2013/14** | **2014/15** | **2015/16** | **2016/17** | **2017/18** | **2018/19** | **2019/20** | **2020/21** | **2021/22** | **2022/23** |
| Overall |  |  |  |  |  |  |  |  |  |  |
| Long-term vaping | 1.6  [1.4–1.8] | 2.5  [2.2–2.7] | 2.9  [2.6–3.2] | 3.3  [3.0–3.5] | 3.5  [3.2–3.8] | 3.2  [2.9–3.5] | 3.9  [3.6–4.2] | 4.9  [4.5–5.2] | 4.9  [4.5–5.3] | 8.6  [7.9–9.3] |
| By vaping frequency |  |  |  |  |  |  |  |  |  |  |
| Long-term daily vaping | 0.9  [0.7–1] | 1.5  [1.3–1.7] | 2.0  [1.8–2.3] | 2.4  [2.2–2.6] | 2.5  [2.3–2.8] | 2.4  [2.1–2.6] | 2.8  [2.5–3.0] | 3.3  [3.0–3.6] | 3.4  [3.0–3.7] | 5.7  [5.2–6.3] |
| Long-term non-daily vaping | 0.6  [0.5–0.7] | 0.8  [0.7–1.0] | 0.7  [0.6–0.8] | 0.6  [0.5–0.7] | 0.7  [0.6–0.9] | 0.6  [0.4–0.7] | 0.7  [0.5–0.8] | 0.9  [0.7–1.0] | 0.8  [0.6–1.0] | 1.3  [1.0–1.5] |
| By main device type used^2^ |  |  |  |  |  |  |  |  |  |  |
| Long-term disposable vaping | - | - | - | 0.1  [0.1–0.2] | 0.1  [0.1–0.2] | 0.1  [0.1–0.2] | 0.1  [0.0–0.1] | 0.1  [0.1–0.2] | 0.7  [0.5–0.8] | 2.9  [2.5–3.3] |
| Long-term refillable vaping | - | - | - | 2.7  [2.5–3.0] | 2.8  [2.5–3.0] | 2.6  [2.3–2.8] | 3.1  [2.8–3.4] | 3.9  [3.6–4.2] | 3.3  [2.9–3.7] | 4.6  [4.1–5.1] |
| Long-term pod vaping | - | - | - | 0.4  [0.3–0.5] | 0.5  [0.4–0.7] | 0.5  [0.4–0.6] | 0.5  [0.4–0.6] | 0.6  [0.5–0.7] | 0.7  [0.6–0.9] | 1.0  [0.7–1.2] |

^1^ Data are weighted proportions, overall (across all participants surveyed between October 2013 and October 2023) and aggregated by survey year (October through September).

^2^ Estimates of prevalence by vaping frequency and main device type used do not sum to the overall prevalence because they do not include participants who responded ‘don’t know’ or who did not provide a response.

^3^ The main device type used was not assessed before July 2016.

##### Table S3. Modelled estimates of changes in the prevalence of long-term vaping among never smokers by age

|  | **Long-term vaping,**  **% [95% CI]**^1^ | |
| --- | --- | --- |
|  | **October 2013** | **October 2023** |
| Age (years)^2^ |  |  |
| 18 | 0.0 [0.0–0.0] | 16.1 [11.1–22.7] |
| 25 | 0.0 [0.0–0.1] | 8.1 [6.3–10.4] |
| 35 | 0.1 [0.0–0.2] | 2.9 [2.0–4.4] |
| 45 | 0.2 [0.1–0.6] | 1.1 [0.6–2.1] |
| 55 | 0.2 [0.1–0.4] | 0.5 [0.3–1.0] |
| 65 | 0.0 [0.0–0.2] | 0.3 [0.1–0.6] |

CI, confidence interval.

^1^ Data for October 2013 and October 2023 are weighted estimates of prevalence in these months from logistic regression with survey month modelled non-linearly using restricted cubic splines (five knots). October 2013 and October 2023 were the first and last months in the time series.

^2^ Note that the model used to derive these estimates included data from participants of all ages, not only those who were aged exactly 18, 25, 35, 45, 55, or 65 years.

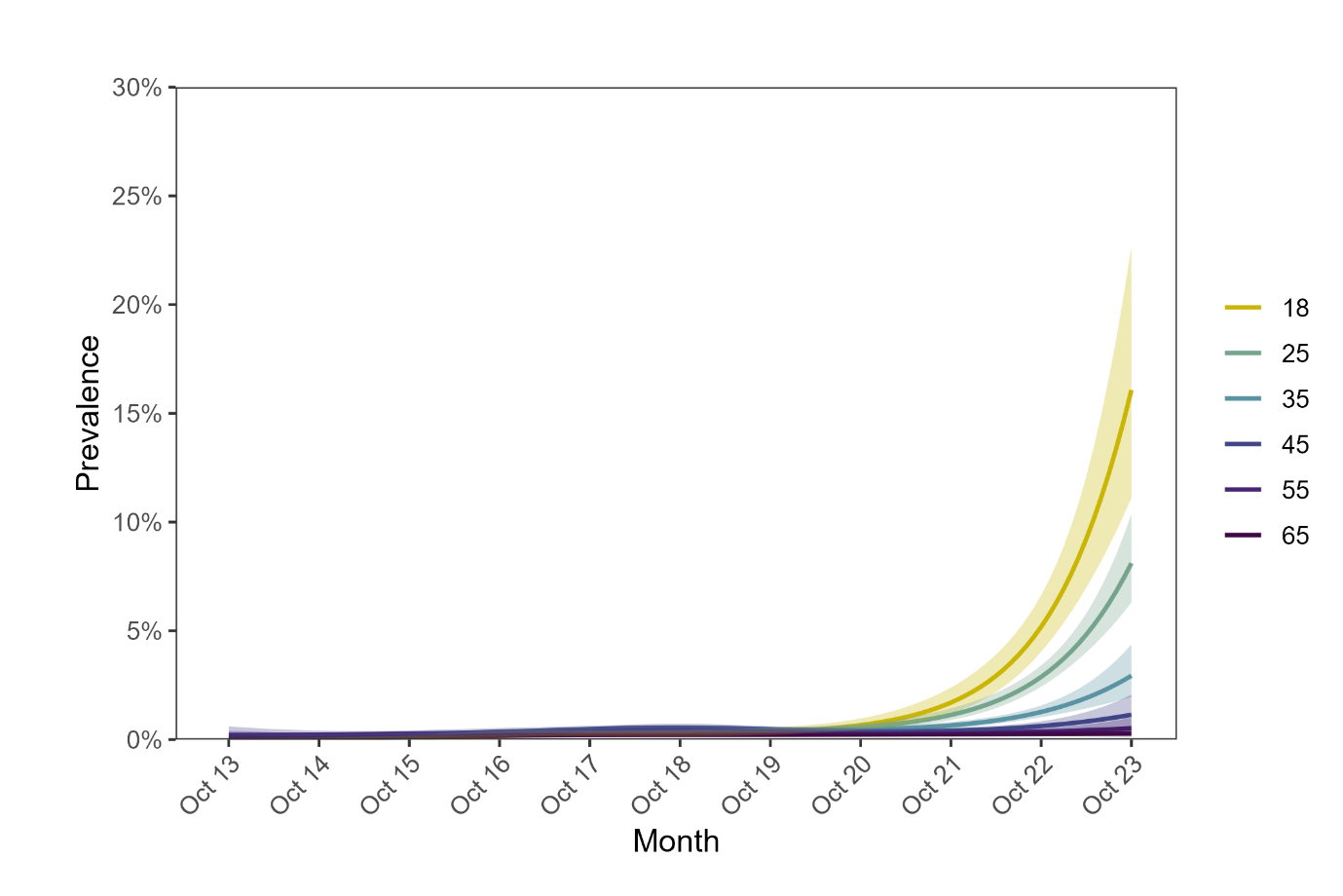

##### Figure S1. Trends in long-term vaping among never smokers by age, October 2013 to October 2023

The figure shows trends in the prevalence of long-term (>6 months) vaping among adults in England who have never regularly smoked, by age. Lines represent modelled weighted prevalence by monthly survey wave, modelled non-linearly using restricted cubic splines (five knots). Shaded bands represent 95% confidence intervals.
